# RSV neutralizing antibodies in dried blood

**DOI:** 10.1101/2023.08.10.23293875

**Authors:** Jonne Terstappen, Eveline M. Delemarre, Anouk Versnel, Joleen T. White, Alexandrine Derrien-Colemyn, Tracy J Ruckwardt, Louis Bont, Natalie I. Mazur

## Abstract

The key correlate of protection of respiratory syncytial virus (RSV) vaccines and monoclonal antibodies (mAb) is virus neutralization, measured using sera obtained through venipuncture. Dried blood obtained with a finger prick can simplify acquisition, processing, storage, and transport in trials, and thereby reduce costs. In this study we aim to validate a low-tech assay to measure RSV neutralization in dried capillary blood. Recovery of mAb from dried blood (volumetric absorptive microsampling) was used to validate the elution method using indirect ELISA. Functional antibodies measured by a neutralization assay were compared between matched serum and dried blood samples from a phase I trial with RSM01, a novel investigational anti-RSV Prefusion F mAb. Hep-2 cells were infected with a serial dilution of sample-virus mixture using RSV-A2-mKate to determine half-maximal inhibitory concentration. Stability of dried blood was evaluated over time and during temperature stress. Functional antibodies in dried blood were highly correlated with serum (R^2^ = 0.98, p < 0.0001). The intra-assay, inter-assay, and inter-operator precision of the assay for dried blood was similar to serum. The function of mAb remained stable for 9 months at room temperature and frozen dried blood samples but lower concentrations showed instability after 6 months. Dried blood samples resisted 48 hours of temperature stress. We demonstrated the feasibility of measuring RSV neutralization using dried blood as an alternative to serum. Measuring antibody function using dried blood is a patient-centered solution that may replace serology testing in trials against RSV or other viruses, such as influenza and SARS-CoV-2.

**Summary points:**

- Neutralizing antibodies against RSV in serum and dried blood from clinical samples are highly correlated.
- Neutralizing antibodies are stable in dried blood for 6 months and can withstand temperature variation.
- Dried blood samples are a patient-centered and logistical solution for vaccine trials in remote areas and low- and lower-middle income countries.

## Introduction

The respiratory syncytial virus (RSV) vaccine and monoclonal antibody (mAb) landscape has rapidly expanded with 16 candidates currently in late phase clinical development [1]. A next-generation, long-acting mAb was approved in 2022 and expected to be commercially available in 2023 [2, 3], soon followed by vaccines for pregnant women and older adults. All late phase vaccines are in development for high income countries due to the high costs, so a delay in implementation is expected in areas with the highest infant RSV burden: low- and lower-middle income countries (LMICs) [4]. RSM01 (an anti-RSV Prefusion (PreF) protein site ø mAb) is intended for infants in LMICs and recently completed a phase 1a trial in healthy adults [5].

Trials largely monitor neutralizing antibody concentrations in serum as a correlate of protection. The functional antibodies are measured by a broad variety of neutralization assays harmonized with an international standard [6]. Blood draws are burdensome for trial participants due to the large amount of blood taken through painful venipunctures that can fail or cause a hematoma. The cost of personnel and equipment, immediate processing, and the necessity of a -80°C cold-chain is resource-intensive and complicates implementation of trials in LMICs. Moreover, venipuncture is difficult to perform in children. Alternatively, dried blood samples can simplify trials, especially in remote areas (Figure 1): one drop of blood is obtained with a finger or heel prick by self-sampling or minimally trained personnel. Samples can be shipped by regular mail without temperature control and stored at -20°C before analysis, while serum samples require clotting and centrifugation before storage or analysis.

**Figure 1.**
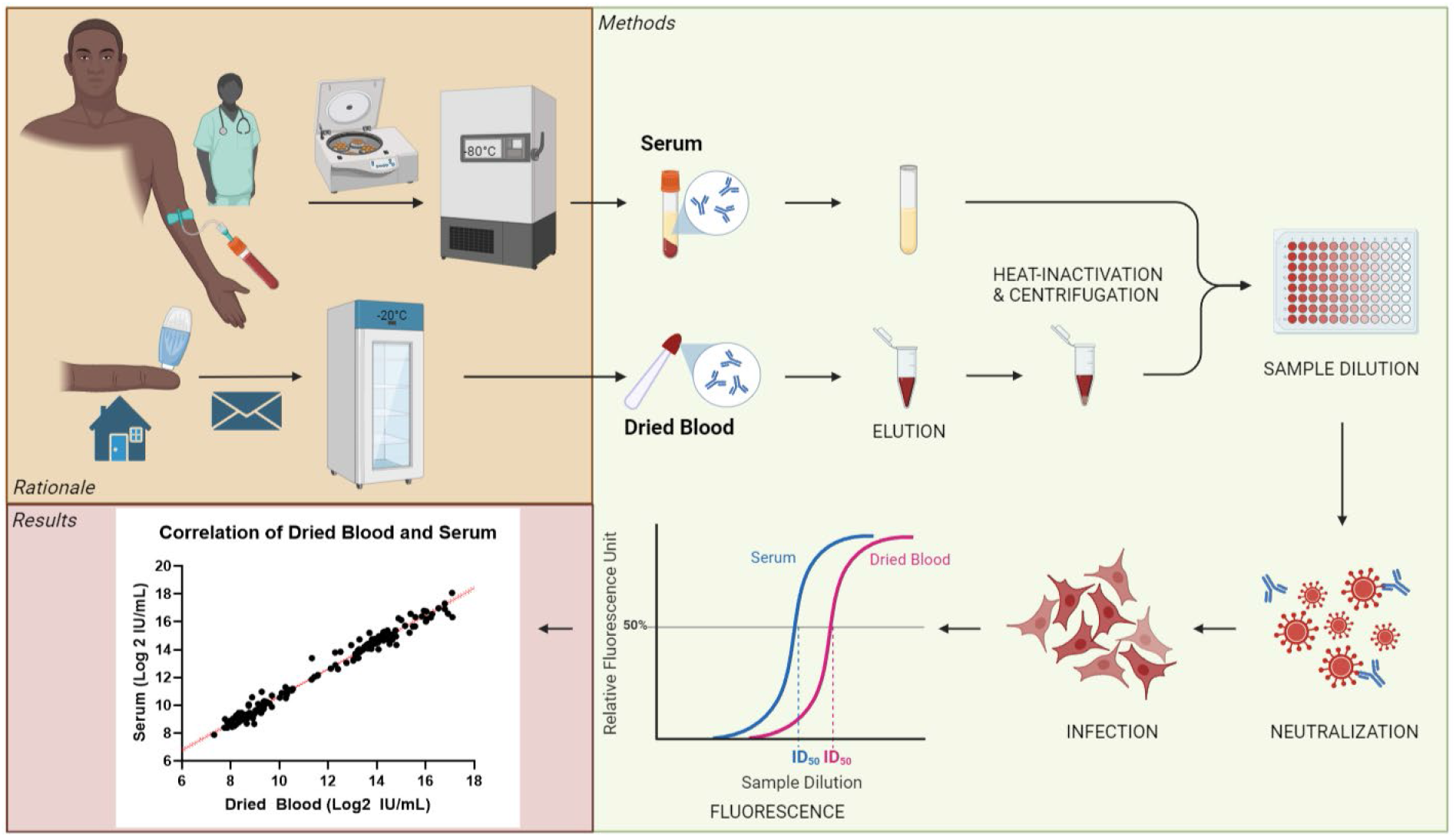
Study rationale and experimental set-up. Trials conventionally monitor neutralizing antibody levels in serum, which requires a professional to draw blood, immediate processing in a lab, and storage and transport at very low temperatures until analysis. Alternatively, dried blood samples can be obtained from a simple finger prick either by non-medical personnel or study participants themselves. Transport through regular post and storage at room temperature or in a regular freezer reduces the necessary resources allowing trials in remote areas. Antibody function was evaluated in dried blood versus serum on the neutralization assay. Abbreviations: ID50, half-maximum inhibitory dilution; mAb, monoclonal antibody; PreF, prefusion protein; RSV, respiratory syncytial virus. This figure was created with Biorender.

Dried blood using filter paper or Mitra^®^ volumetric absorptive microsamples (VAMS^®^) is validated to measure mAb concentrations measured by ELISA [7], but neutralizing capacity of antibodies after drying of blood is less well-established. Five studies compared neutralizing antibodies in dried blood spots (DBS) on filter paper or hemaPEN^®^ with plasma or serum for SARS-CoV-2, rabies and human papilloma virus [8-12]. Samples sizes were small (range 7-48 matched samples, total 128 pairs) with varying correlations (R^2^ between 0.51-0.99). This study aimed to determine if anti-RSV antibodies retain their function in dried blood using VAMS^®^ devices in a large sample size using clinical samples from the first-in-human trial with RSM01. Secondly, we validated the use of dried blood compared to serum using a simple tool – a reporter virus-based neutralization assay – to measure neutralizing antibodies (nAb) against RSV.

## Methods

### Sample handling and preparation

Individual samples for validation of the assay were obtained from healthy adults from the University Medical Center Utrecht healthy donor service. Serum and whole blood were unaltered or spiked with 1-100 μg/mL RSM01, a mAb against RSV PreF site ø (Bill & Melinda Gates Medical Research Institute). After 30 minutes on a rocker to mix, Mitra^®^ 20 μl VAMS^®^ (Trajan Scientific, Neoteryx Clamshell, 20004) were touched to the whole blood sample surface to wick up the fixed volume [13] and left to dry for 3 hours at room temperature (RT). Samples were used immediately or stored at -80°C or higher temperatures (RT or -20°C) for stability assessments.

Clinical samples at matched timepoints (Day 1 pre-dose, Day 91, Day 151) were collected from 56 participants in Clinical Study GATES-MRI-RSM01-101 (NCT05118386): by venipuncture blood in serum separator tubes processed to serum or as finger-prick via lancet followed by touching the Mitra^®^ 20 μl VAMS^®^ to the blood drop to wick up the fixed volume. Of note, the adult participants in this study were expected to have pre-existing RSV neutralizing antibodies from natural infection. Samples were stored at - 80°C. Prior to analysis on the neutralization assay, dried blood samples were thawed for 30 minutes before opening the air-tight bag to prevent condensation.

### Dried blood elution for antibody recovery

Dried blood was rehydrated with a PBS-only elution method: one microsampler tip was transferred to a 2 mL Eppendorf tube with 200 μL 1×PBS and incubated overnight at RT on a shaker at 300 rpm. The PBS-only elution method was compared to a validated elution method, which incubates microsampler tips in 400 μL of 1×PBS + 1% BSA + 0.5% Tween20 at 4°C overnight as previously published [14]. Antibody recovery was measured on an RSV PreF ELISA (Supplementary Methods). The elimination of Tween from the elution buffer is important to enable use in cell-based assays such as this RSV neutralization assay. The elimination of BSA from the elution buffer is important to enable use in resource-limited regions where acquisition of animal-based reagents is limited.

%Absolute recovery is the percentage of measured concentration divided by spiked concentration. %Recovery is calculated as the (measured concentrations of the spiked sample – unaltered sample)/measured concentration of the spiked standard diluent*100%. Elution efficiency was defined as the %recovery with preferred PBS-only elution method divided by the %recovery with the validated elution method. Boundaries for acceptance were set at 80-120%.

### Neutralization assay

Neutralizing capacity of samples was tested using a neutralization assay described previously [15, 16]. Briefly, Hep-2 cells (ATCC CCL-23) were seeded at a concentration of 0.5 × 10^6^ cells/well in 384-well black optical bottom plates (Thermo Scientific, 142761) and incubated for > 4 hours. Dried blood eluate and serum were heat-inactivated at 56°C for 30 minutes. Dried blood eluate was then centrifuged at 15.000g for 10 minutes to remove cell debris. Serum and dried blood eluate (1:10during elution) were serially diluted in 3-fold from starting dilution 1:5 or 1:2 (total dilution 1:20), respectively, in Dulbecco’s modified Eagle’s Medium containing 10% FCS (Sigma High Glucose, D6429 or Gibco Glutamax, 31966-021). Recombinant mKate-RSV-A2 (2.23×10^6^ TCID50/mL) [17, 18] was added 1:1 to the sample dilution series and incubated for 1 hour at 37°C before addition of 50 μL of sample-virus mixture to the adherent Hep-2 cells. After 26 hours incubation at 37°C, we recorded relative fluorescence units (RFU) using excitation at 584 nm and emission at 620 nm (FLUOstar OMEGA, BMG Labtech). The neutralization curve and 50% inhibitory concentration or dilution (IC50 or ID50, respectively) of samples were analyzed using a log (inhibitor) vs response 4-parameter nonlinear regression curve in GraphPad Prism version 8.3 (GraphPad Software Inc., San Diego, CA, USA). International units (IU) were calculated according to manufacturer’s instruction (Supplementary Methods).

### Sample stability

Stability of mAb in dried blood was tested with dried blood and serum samples spiked with 10 or 100 μg/mL RSM01. Dried blood samples were stored in generic zip-lock bags including silica gel and humidity cards.

Long term stability was evaluated because the opportunity to delay sample processing could ease logistical challenges of trials in remote areas. Dried blood was stored at -20°C and RT while serum aliquots were stored at -80°C. Storage temperature was logged to ensure correct storage conditions. Samples were measured on the neutralization assay at baseline (1 day after sample preparation) and at seven follow up time-points over one year. Stability was defined as less than 30% difference in ID50 per time-point compared to baseline ID50.

Short term stability with a stress test was evaluated to simulate potential varying storage conditions during sample transport. Spiked dried blood samples were stored at -20°C and underwent temperature stress at various degrees (4°C, RT, 37°C or 45°C) for a total of 48 hours before returning to - 20°C storage. Stability was defined as less than 30% difference in ID50 between stressed samples and samples that were not stressed but remained at -20°C.

### Statistical Analysis

Critical reagents were determined by paired t-test of linear ID50 values of the same sample measured with two different reagents (IBM SPSS Statistics, version 26.0.0.1). Precision was calculated as the %CV of duplicates of the sample (intra-assay precision), %CV of the linear ID50 of several runs between days (inter-assay precision) or the %CV of the same run by two operators (inter-operator precision). Dried blood and serum from the RSM01 Phase 1 clinical trial were compared using simple linear regression (IBM SPSS Statistics, version 26.0.0.1).

### Ethical considerations

Informed consent was obtained from healthy donors and study participants. The healthy donor service was approved by the Ethical Committee of Biobanks (study number 18-774) and medical ethical committee (07-125/O). The RSM01 phase 1 clinical trial (NCT05118386) was conducted in accordance with the United Department of Health and Human Services guidelines and the ethical standards of the Helsinki Declaration (2013).

## Results

### Validation of Elution Method

We first validated our PBS-only elution method against an existing validated elution method [14] by testing PreF binding by samples recovered from dried blood with or without spiked RSM01. Acceptance limits were met by 90% of the samples for both elution efficiency and absolute recovery (Figure 2A and B). Recovery was consistent over two days with a mean CV of 5.4% ± 4.3 SD (Figure 2C). Based on our findings, our PBS-only method was considered non-inferior to the validated method for recovery of RSV PreF-binding antibody.

**Figure 2.**
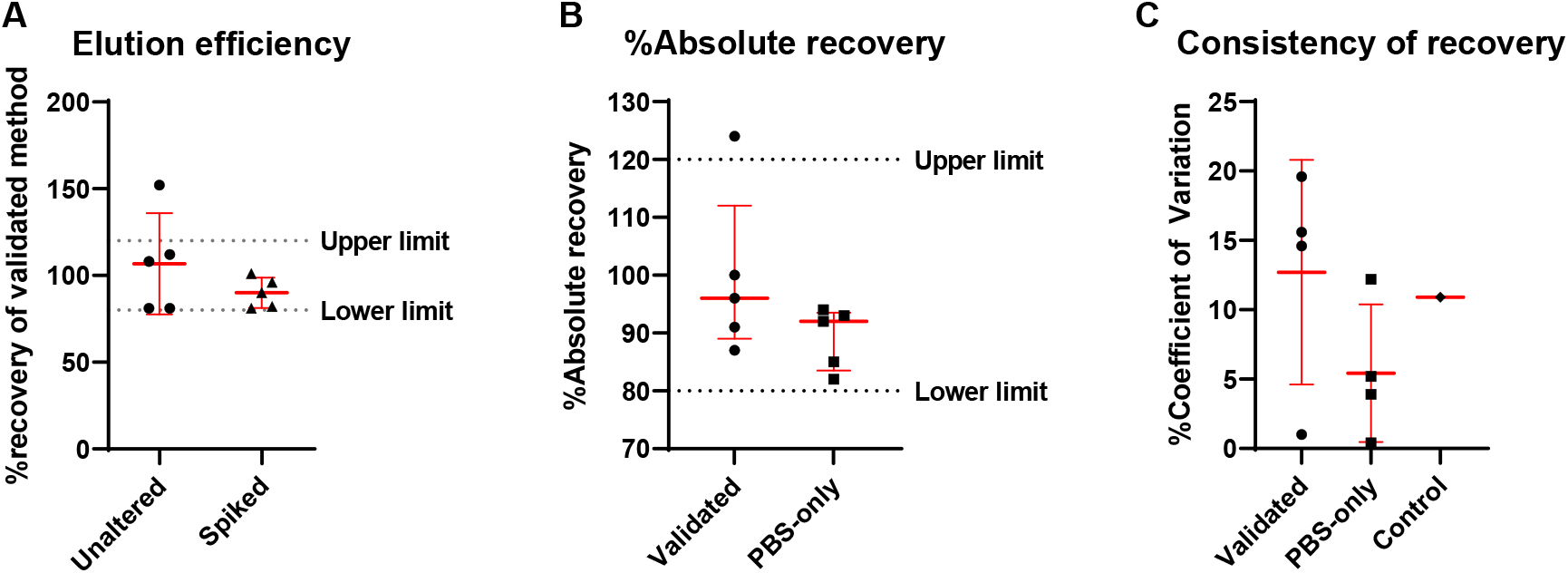
Comparison of two elution methods on dried blood samples unaltered or spiked with 100 μg/mL RSM01 in healthy donor peripheral blood measured by RSV preF ELISA. Comparison of validated vs PBS-only elution method (n = 5 samples from 3 healthy donors) **A**| Elution efficiency was defined as the %recovery with preferred PBS-only elution method divided by the %recovery with the validated elution method (mean ± SD). **B**| %absolute recovery per elution method (mean ± SD). %Absolute recovery is the percentage of measured concentration divided by spiked concentration. %Recovery is calculated as the (measured concentrations of the spiked sample – unaltered sample)/measured concentration of the spiked standard diluent*100%. **C**| Consistency of recovery expressed as %CV between two days (mean ± SD). RSM01 100 μg/mL in PBS served as control. %CV is the standard deviation divided by the mean ^*^ 100%. Abbreviations: CV, coefficient of variance; ELISA, enzyme linked immunosorbent assay; PreF, Prefusion glycoprotein; RSV, respiratory syncytial virus; SD, standard deviation.

### Validation of Dried Blood on RSV Neutralization Assay

#### Validation of RSV Neutralization Assay

We determined the critical reagents for the RSV neutralization assay. Two virus batches cultured from the same master stock appeared to affect the ID50, but it did not achieve significance (p=0.057; Figure 3A). Two brands of medium (Sigma and Gibco) for cell culture affected the ID50 of samples significantly (p = 0.009; Figure 3B), while it had no effect when used for the dilutions (p = 0.2161; Figure 3C). Based on this data, culture medium and virus batch are considered critical reagents that require characterization, while the dilution medium is not.

**Figure 3.**
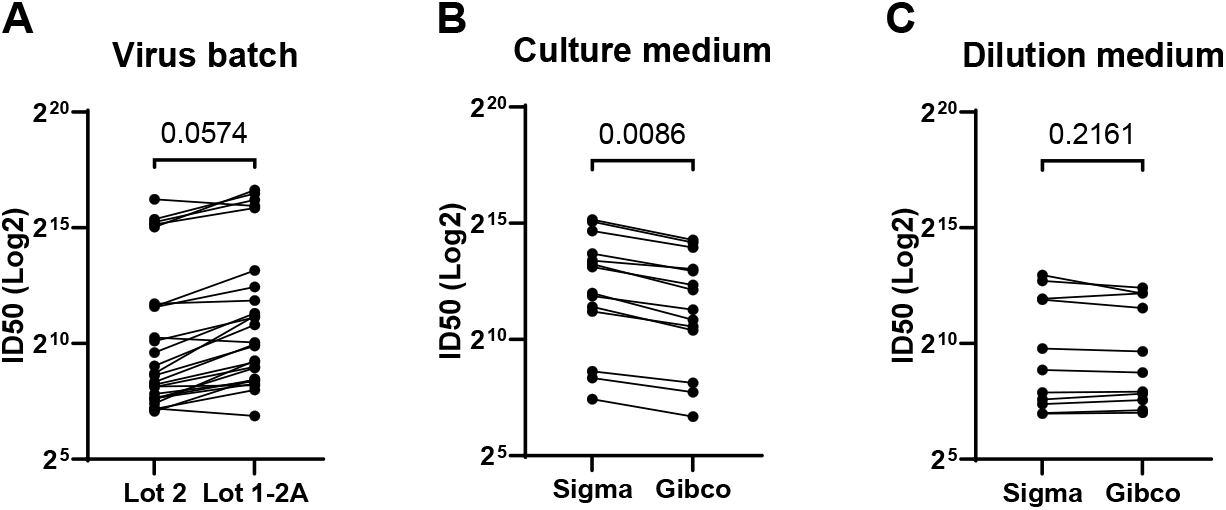
Characterization of critical reagents for RSV neutralization assay on dried blood. **A**| Comparison of assay performance with two virus batches. **B**| Comparison of assay performance with two brands of culture medium, a critical reagent. **C**| Comparison of assay performance with old versus new dilution medium, a non-critical reagent. Significance determined by paired t-test. Abbreviation: ID50, half-maximal inhibitory dilution.

A range of concentrations of RSM01 in PBS was evaluated to determine the limits of detection on the neutralization assay. We found a wide dynamic range of the assay, enabling measurement of RSM01 in concentrations ranging from 1 μg/mL up to more than 10 mg/mL (Figure 4).

**Figure 4.**
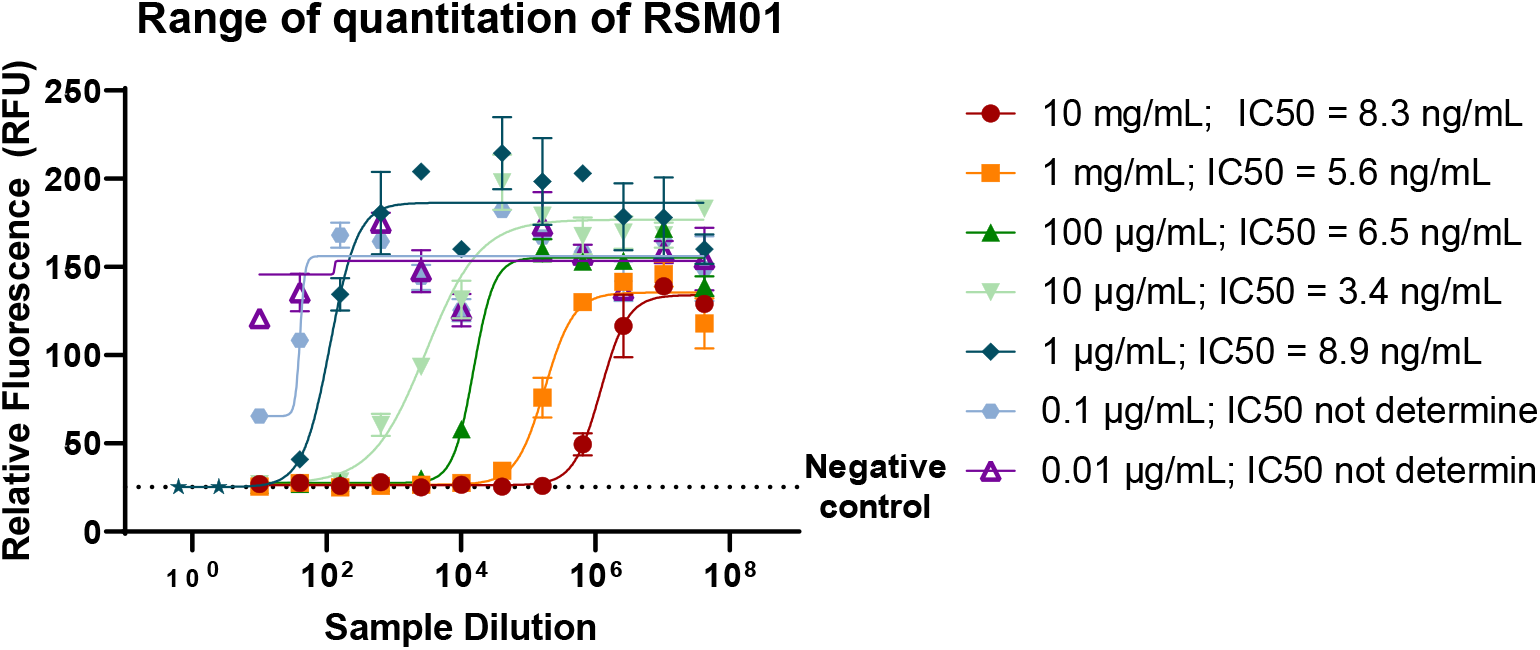
Characterization of limits of detection of RSV neutralization assay. Dynamic range of quantitation of RSM01 in PBS on the neutralization assay. Error bars resemble technical duplicates. The lowest two concentrations did not qualify the assay quality control criteria and IC50 values could not be determined. Abbreviation: IC50, half-maximal inhibitory concentration.

Precision of the RSV neutralization assay with dried blood (≤28.1%CV) was non-inferior to serum samples (≤34.0%CV) (Supplemental Table 1).

#### Stability of Dried Blood

We evaluated long term storage of dried blood samples compared to serum at room temperature (RT) and -20°C. Dried blood with 100 μg/mL (high) or 10 μg/mL (low) concentrations of RSM01 retain stable functionality compared to serum (Figure 5A). An outlier of serum 100 μg/mL at 1 month storage is likely an artifact due to faulty measurement (as the is unlikely to be an increase in concentration and because subsequent points show stability). What appears as loss of neutralizing capacity after 1 month storage could also be caused by faulty baseline measurement. Therefore, we calculated an adjusted baseline using all available data of heathy donor samples spiked with 100 μg/mL RSM01 (n = 18 of 3 healthy donors) or 10 μg/mL (n = 11 of 2 healthy donors) as natural immunity is negligible when spiked with these concentrations of RSM01. Antibodies at higher concentrations retain their function for 9 months in frozen and RT dried blood as well as in frozen serum, but at lower concentrations mAb the loss of function is already observed after 6 months storage (Figure 5B). Long term storage (>6m) of dried blood at RT showed discoloration that increased over time from maroon to brown-grey. Humidity was less than 10% over time in dried blood samples at either storage condition over time.

**Figure 5.**
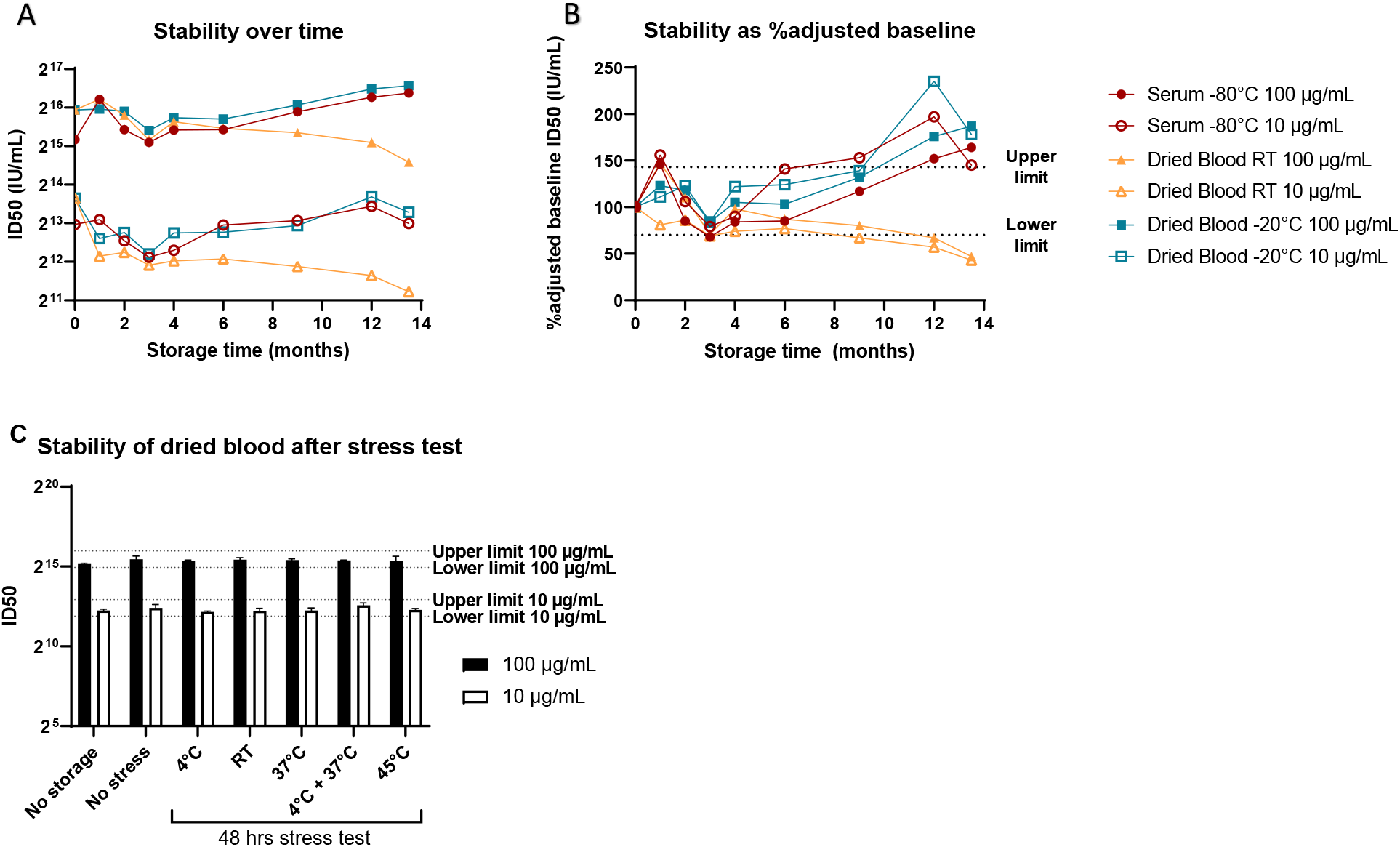
Stability of antibody function in dried blood samples compared to serum samples. Stability of functional antibodies in dried blood stored at room temperature and -20°C compared to serum stored at -80°C (baseline n = 2, other timepoints n = 1). Preset upper and lower limit of acceptance are 143% and 70%, respectively (dotted lines). **A**| Neutralizing capacity after long-term storage expressed as ID50 in IU/mL. **B**| Neutralizing capacity of dried blood expressed as the percentage of adjusted baseline neutralizing capacity (ID50 in IU/mL). Adjusted baseline was calculated as the geometric mean titer of all available data of healthy donor blood spiked with 100 μg/mL (n = 18) or 10 μg/mL (n = 11) excluding the long term stability data. **C**| Dried blood sample stability after short-term storage at -20°C with 48hrs stress test at various temperatures before returning to storage at -20°C. Upper and lower limit of acceptance are based on stable storage at -20°C (no stress) condition. Abbreviations: ID50, half-maximum inhibitory dilution; IU, international units; RT, room temperature.

Short term storage at -20°C followed by 48 hours of temperature stress did not induce sample instability at either spiked concentration (Figure 5D). Dried blood samples were kept at humidity <10% throughout the temperature stress and the samples at higher temperatures > 37°C showed brown-grey discoloration similar to that observed at > 6 months storage at RT.

#### Comparison of functional antibodies in dried blood and serum on RSV Neutralization Assay

We compared the neutralizing activity in dried blood eluate to serum at three different timepoints for 56 participants of a Phase 1 clinical trial evaluating RMS01 in healthy adults. Neutralizing activity in dried blood samples correlated strongly with serum neutralization (R^2^ = 0.98, p < 0.0001, n = 165 matched samples, Figure 6A). The duration of storage between sample collection and measurement did not have an effect on the ratio between the matched dried blood and serum samples (slope ≈ 0, mean ratio 0.69, range storage time 29-280 days, Figure 6B).

**Figure 6.**
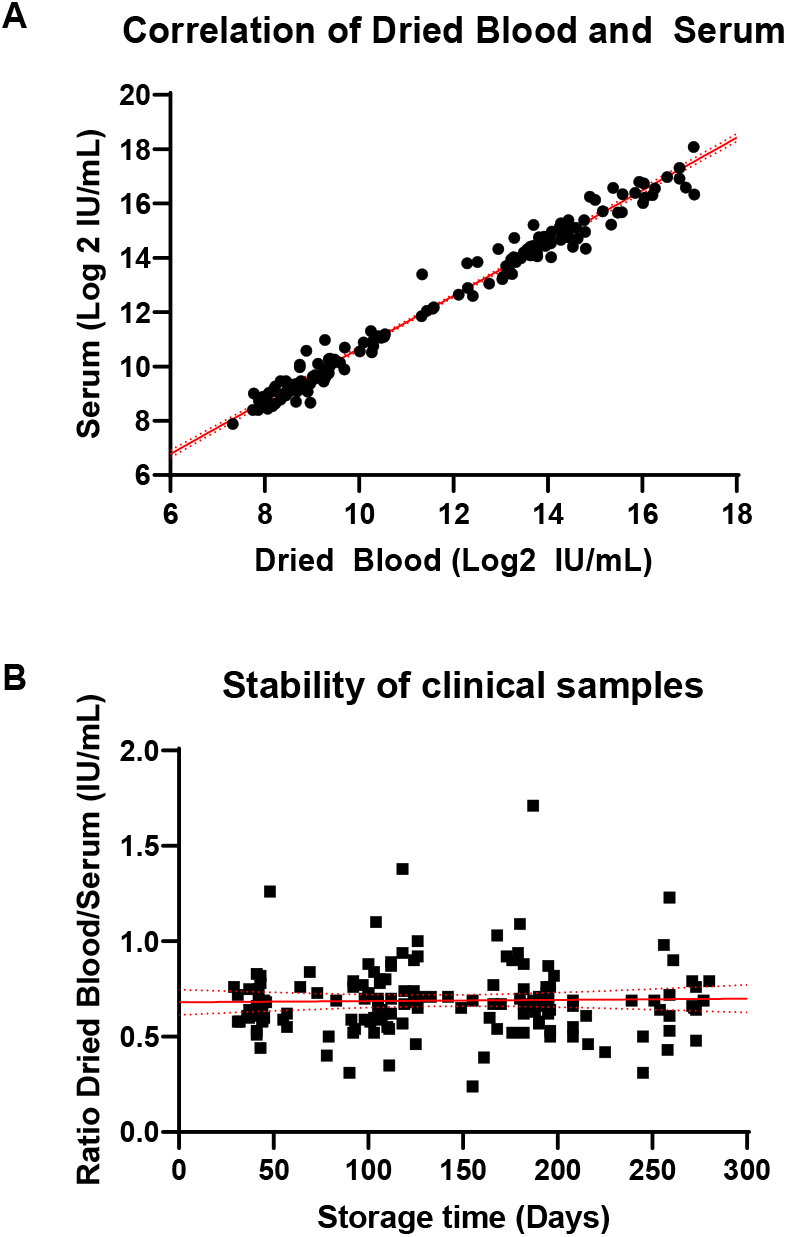
Antibody function in dried blood samples compared to serum samples. **A**| Simple linear regression of Log2 ID50 values of RSM01 Phase 1 clinical trial serum and dried blood samples in IU/mL at three time points (R^2^ = 0.981, n = 165 matched samples) with 95% confidence interval (red dotted lines). **B**| Simple linear regression of the ratio between dried blood and serum ID50 in IU/mL (R^2^ < 0.001, slope ≈ 0) with 95% confidence interval (red dotted lines). Abbreviations: ID50, half-maximum inhibitory dilution; IU, international units.

## Discussion

In this study, we show for the first time that antibodies stored in dried blood retain their neutralizing capacity against RSV and we validated an RSV neutralization assay for reconstituted dried blood. The strong correlation between the two sample types collected in a clinical trial (R^2^ = 0.98) indicates that dried blood could be used to measure RSV neutralizing antibodies. Dried blood samples are highly stable compared to serum, for at least 9 months storage at -20°C and 6 months at RT. Short term temperature stress did not induce instability.

This study is the first to use dried blood samples to measure anti-RSV antibody function. To date, antibody concentration, but not function, has repeatedly been studied in dried blood. The concentrations of therapeutic mAbs are reliably recovered from VAMS [7]. Anti-SARS-Cov-2 antibody titers in VAMS highly correlate with serum suggesting that VAMS is a valid method to evaluate antibody titers [14, 19]. The varying correlation and precision in previous head-to-head comparisons of neutralizing antibodies [8-12] using DBS on filter paper instead of VAMS could be explained by the smaller sample sizes or by the hematocrit effect. This effect is a well-known limitation of DBS where the hematocrit-dependent blood flow through the paper complicates reproducible recovery of the serum fraction containing the antibodies of interest when punching a fixed area of the card [20]. VAMS circumvents these limitations by ensuring homogenous sample uptake and the entire sample is eluted allowing more consistent recovery [20]. Other devices that control total volume are anticipated to have acceptable precision similar to that shown here with VAMS.

Antibody function is widely assumed to be stable over time in serum but it is less known if the same is true for dried blood. According to a systematic review, DBS are most stable with the least variation at - 20°C and -70°C regardless of the intended measurements [21]. No clear decrease in mAb concentration is observed in VAMS up to 1 month storage at RT and 4°C [7] and HIV antibodies are stable when stored for 6 weeks at 4°C, -20°C or -70°C in DBS [22]. We showed that functional antibody in dried blood stored at RT and -20°C is stable compared to adjusted baseline up to at least 6 months storage. However, the neutralizing capacity drops between baseline and month 1 raw data (Figure 5A). We believe that limitations of our study design underestimate stability when comparing to a single baseline measurement at T=0, which was unexpectedly high. Therefore, we calculated a theoretical baseline using various healthy donors to correct this limitation. After short term storage in the stress test no loss in neutralizing capacity was observed, supporting the potential faulty baseline measurement in the long-term stability test (Figure 5C). Moreover, dried blood samples neutralizing capacity plateaus between months 1 and 9 (Figure 5A) concluding that the samples are most likely stable. Our data suggests that -20°C is more suitable for storage > 6 months but shorter storage is also acceptable at RT. These findings need to be confirmed in repeat experiments. We observe that 48 hours of temperature variation – e.g. during shipments of samples – is acceptable in terms of sample stability, although IgG concentration (and like neutralizing capacity) in DBS on filter paper was shown to decrease drastically after 7 days storage at 45°C [23].

This study has several strengths and limitations. The neutralization assay is sensitive to several reagents due to the live cells and virus, which showcases the importance of same batch measurements and the use of assay controls. The enrichment of the media source might affect viral infection and/or replication and thus influence the read-out. The in-house controls allowed for conversion of the results to international units to compare with other RSV neutralization data worldwide and correct for batch variations during long studies. A limitation is that we have validated the assay on dried blood only from healthy adults. Considering the intended use in infants – with maturing immune systems and possibly with comorbidities - we need to continue validation on blood from healthy infants and those with comorbidities such as prematurity, coinfection, or immune deficiencies. Lastly, as assay precision did not meet the 30%CV target for serum samples, it suggests that the ±30% target may have been overly conservative for a cell-based method with two living components, cells and virus.

We demonstrated RSV neutralization on dried blood as a patient-centered solution that may replace serology testing in trials in LMICs and potentially be used as a tool to monitor protection against RSV globally. Our study has implications for the future of vaccine and mAb development against RSV and other respiratory viruses such as influenza and SARS-CoV-2 as blood draws can be challenging in remote areas, especially in young children. One obstacle for worldwide vaccine and mAb access is globally representative trials that need to take place in countries with the highest disease burden because efficacy can differ between regions [24]. The potential to use dried blood obtained with a simple finger prick can reduce logistical burden of trained phlebotomists and cold chain requirements and thereby reduce cost and complexity. We plan to validate the assay in a LMIC setting as proof-of-concept that sample analysis is independent of an advanced immunology lab in a high-income country.

## Contributions

JT, JTW, LB, and NIM were involved in the design and plan for this study. JT, EMD, and AV were involved in the data collection and analysis. TJR and ADC offered methodology transfer, materials, and support during study conduction. JT, LB and NIM were involved quality assessment and contributed to the writing of the manuscript, in collaboration with all co-authors.

## Funding

This work was supported, in whole or in part, by the Bill & Melinda Gates Foundation [grant number INV-008522]. Under the grant conditions of the Foundation, a Creative Commons Attribution 4.0 Generic License has already been assigned to the Author Accepted Manuscript version that might arise from this submission.

## Data availability

Clinical data not publicly available. On reasonable request to the corresponding author, the validation data can be shared.

## Declaration of interest

UMCU has received major funding (>€100,000 per industrial partner) for investigator initiated studies from AbbVie, MedImmune, AstraZeneca, Sanofi, Janssen, Pfizer, MSD and MeMed Diagnostics, the Bill and Melinda Gates Foundation, Bill & Melinda Gates Medical Research Institute and the Dutch Lung Foundation. UMCU has received major funding as part of the public private partnership IMI-funded RESCEU and PROMISE projects with partners GSK, Novavax, Janssen, AstraZeneca, Pfizer and Sanofi. UMCU has received major funding by Julius Clinical for participating in clinical studies sponsored by MedImmune and Pfizer. UMCU received minor funding (€1,000-25,000 per industrial partner) for consultation and invited lectures by AbbVie, MedImmune, Ablynx, Bavaria Nordic, MabXience, GSK, Novavax, Pfizer, Moderna, Astrazeneca, MSD, Sanofi, Janssen. LJB and NIM have regular interaction with pharmaceutical and other industrial partners. They have not received personal fees or other personal benefits. LJB is the founding chairman of the ReSViNET Foundation. JTW is an employee at the Bill & Melinda Gates Medical Research Institute. ADC and TR are supported by funding from intramural NIAID.

## Notes

### Competing Interest Statement

UMCU has received major funding (>100,000 euro per industrial partner) for investigator initiated studies from AbbVie, MedImmune, AstraZeneca, Sanofi, Janssen, Pfizer, MSD and MeMed Diagnostics, the Bill and Melinda Gates Foundation, Bill & Melinda Gates Medical Research Institute and the Dutch Lung Foundation. UMCU has received major funding as part of the public private partnership IMI-funded RESCEU and PROMISE projects with partners GSK, Novavax, Janssen, AstraZeneca, Pfizer and Sanofi. UMCU has received major funding by Julius Clinical for participating in clinical studies sponsored by MedImmune and Pfizer. UMCU received minor funding (1,000-25,000 euro per industrial partner) for consultation and invited lectures by AbbVie, MedImmune, Ablynx, Bavaria Nordic, MabXience, GSK, Novavax, Pfizer, Moderna, Astrazeneca, MSD, Sanofi, Janssen. LJB and NIM have regular interaction with pharmaceutical and other industrial partners. They have not received personal fees or other personal benefits. LJB is the founding chairman of the ReSViNET Foundation. JTW is an employee at the Bill & Melinda Gates Medical Research Institute. ADC and TR are supported by funding from intramural NIAID.

### Author Declarations

TCBio and METC NedMec of University Medical Center Utrecht gave ethical approval for this work. Ethics committee of Bill & Melinda Gates Medical Research Institute gave ethical approval for this work.

