## Supplementary Methods; Supplementary Table 1 for "RSV neutralizing antibodies in dried blood"

Terstappen et al.

### Supplementary Material

#### Supplementary methods

##### RSV PreF Enzyme Linked Immunosorbent Assay (ELISA)

Nunc MaxiSorp 96-well plates (ThermoFischer Scientific, 442404) were coated overnight at 4°C with 1 µg/mL RSV stabilized PreF (DsCav-1) protein. Plates were washed with PBS containing 0.05% Tween-20 (PBS-T) in between incubation steps and blocked with PBS-T + 1% BSA for 1 hour at RT. The standard curve (0.24-250 ng/mL RSM01) and samples (1000-32000x diluted) were added in duplicate to incubate for 2 hours. The detection antibody (goat anti-human IgG-HRP; Jackson ImmunoResearch; 109-035-098) was added for 1 hour at RT followed by TMB (Biolegend, 421101) until color development was stopped with 1M H2SO4. The absorbance was measured at 415 nm (Spectramax M series, Company Molecular Devices). Sample were quantified using a 4-parameter logistic regression curve of log(concentration) with GraphPad Prism version 8.3 (GraphPad Software Inc., San Diego, CA, USA). Duplicates were accepted if the coefficient of variation (CV) was below 15%.

##### Neutralization assay

We created an in-house low positive control (LPC) by identifying four healthy donors with low neutralizing capacity and a high positive control (HPC) by selecting seven serum samples from our RSV biobank with known nAb titers. Pooled serum was mixed on a roll bank at 4°C overnight before aliquots were stored at -80°C. HPC and LPC were included in all neutralization assay plates as controls.

To standardize neutralization data to the First International Standard (IS) for Antiserum to RSV (NIBSC code 16/284) our in-house HPC and LPC were run simultaneously to the World Health Organization (WHO) International Standard (IS) [17]. Assays were run on three days by two operators for a total of 12 runs to assign critical reagent specific international units (IU) to our HPC/LPC per manufacturer's instructions. The ratios of (IU/mL) / (GMT of HPC or LPC of all runs) were used to generate a conversion factor of 2.01 for Sigma DMEM and 2.75 for Gibco DMEM.

Quality control (QC) criteria for the neutralization assay were adapted from Sarzotti-Kelsoe et al., 2014 [20]: (1) average RFU of virus control > 2x average RFU cell control; (2) CV of RFU in virus control wells < 25%; (3) %CV of duplicate wells of sample dilution < 15%; (4) neutralized plateau within 30% of the average of the cell control wells: cell only RFU values were added as data points (maximum of 3 data points on plateau) to correct the curve in case of < 3 points on the neutralized plateau (with mAb testing) or when the linear section of the curve was present with one point below the expected 50% neutralization (with polyclonal sample testing); (5) positive control titers had to be within 2-fold of the mean of the previous values for that particular critical reagent combination. Data that failed the QC criteria were excluded from analysis (include the duplicate resulting in the highest $R^2$ unless if the curve becomes non-sigmoidal).

#### Supplementary data

Supplemental Table 1| **Precision of the RSV neutralization assay to measure natural immunity and spiked serum and dried blood samples.**

| | **Intra-assay[1]** | **Inter-assay** | **Inter-operator** |
|---|---|---|---|
| Serum | | | |
| Natural immunity | 3.2 (1.4-6.0; n=30) | 17.3 (11.8-29.7; n=18) | 16.1 (4.4-23.8; n=19) |
| Spiked | 5.1 (2.3-8.5; n=12) | 19.6 (12.9-22.1; n=5) | 34.0 (8.7-49.8; n=6) |
| Dried blood | | | |
| Natural immunity | 7.3 (3.2-12.5; n=30) | 22.0 (6.6-34.7; n=13) | 18.5 (9.8-32.8; n=11) |
| Spiked dried | 3.5 (1.6-5.9; n=12) | 28.1 (25.5-32.5; n=4) | 16.6 (7.6-24.5; n=5) |

Precision is expressed as the median %CV (IQR; amount of unique samples). Samples were spiked with 1, 10 or 100 µg/mL RSM01 in serum or whole blood before VAMS preparation. [1]The duplicates of 12 dilutions per unique sample are taken along in intra-assay calculation. Abbreviations: CV, coefficient of variance; IQR, inter-quartile range.
